# An Actor-Critic Reinforcement Learning Framework for Variant Evidence Interpretation

**DOI:** 10.1101/2025.03.14.25323954

**Authors:** Dylan Lawless

**Author notes:** Addresses for correspondence.

## Abstract

We present a reinforcement learning (RL) framework that uses established genomic metrics – such as the GuRu score, variant/gene risk priors, and population frequency – to estimate the probability of observing a given genetic variant in disease. Importantly, our approach does not directly predict variant pathogenicity; instead, it quantifies the cumulative evidence supporting a variant’s clinical observability within a Bayesian context. Using simulated genetic data with a range of variability and label noise, we systematically evaluated the actor-critic algorithm performance across multiple scenarios, employing metrics including receiver operating characteristic (ROC) curves, area under the curve (AUC), calibration plots, and learning dynamics. Results indicate predictive accuracy and effective learning, demonstrating RL’s potential as a practical tool for genomic variant interpretation, setting the stage for integration into a broader Bayesian classification framework.

## 1 Introduction

Precise interpretation of genetic variants remains a central challenge in precision medicine, significantly impacting clinical decision-making and patient care. Differentiating pathogenic from benign variants accurately is essential for genetic diagnostics. Standard classification methods often face limitations due to incomplete or uncertain annotations, motivating the exploration of adaptive machine learning techniques. To address these challenges, we simulate genomic data that captures the main target features of evidence underlying variant interpretation and the inherent uncertainties encountered in clinical settings.

Qualifying variant (QV)s represent genomic alterations identified through stringent criteria in genomic processing pipelines and form the foundation for calculating the GuRu score. In our simulation, each QV is assigned a GuRu score based on the cumulative tally of effect evidence, comprising functional assays, computational predictions, and clinical observations, thereby reflecting the strength of evidence for a variant’s potential pathogenicity. The selection and classification of these QVs adhere to established best practices in variant reporting and analysis (1–5), as well as standardised workflows (6–8). Furthermore, population allele frequencies from databases such as gnomAD (9) inform the estimation of variant probabilities, while resources like ClinVar (10) and ClinGen (11) provide clinical evidence that reinforces the interpretation of variant impact and associated phenotypes. Since this dataset is inherently noisey, we begin by substituting with simpler synthetic data, laying the groundwork for future application of these methods to empirical genomic datasets.

RL provides an appealing alternative to traditional supervised methods by utilising evaluative rather than instructive feedback. Instead of explicitly labelled outcomes, RL algorithms receive scalar rewards reflecting decision quality, thereby naturally balancing exploration of uncertain genomic space and exploitation of known information. Inspired by the classical *k*-armed bandit problem, our approach adapts an actor-critic algorithm to genomic data, using key features already available clinically – such as the GuRu score, gene risk categorisation, and population allele frequency – to classify the probability of a variant being assigned as pathogenic based on the existing evidence. This is subtly but importantly distinct form assigning pathogenicity itself, which is a separate task.

We recall the actor-critic architecture of the pole-balancing control problem as described by Barto et al. (12, 13). In that work, a two-component adaptive system was introduced, comprising an adaptive critic element (ASE) (associative search element) and an associative search element (ACE) (adaptive critic element), designed to solve the challenging pole-balancing control problem without any prior knowledge of the system dynamics. The system learns exclusively from sparse, delayed failure signals. Specifically, the ASE employs a stochastic, reinforcement-based update rule given by

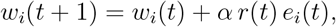

where *w*_*i*_(*t*) denotes the weight of the *i*th input connection at time *t, α* is the actor’s learning rate, *r*(*t*) is the reinforcement signal provided by the environment at time *t*, and *e*_*i*_(*t*) is the eligibility trace capturing the recent history of activity for that connection. The eligibility trace itself decays over time according to

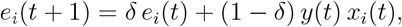

with *δ* representing the decay factor, *y*(*t*) the output of the system at time *t*, and *x*_*i*_(*t*) the *i*th component of the input vector. Meanwhile, the critic’s weight for the *i*th input is denoted by *v*_*i*_(*t*) and is updated via

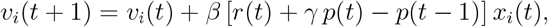

where *β* is the critic’s learning rate, *γ* is the discount factor determining the influence of future rewards, and *p*(*t*) is the critic’s current prediction of the cumulative future reward. This update rule allows the critic to refine its predictions of long-term rewards by comparing the current prediction with the observed reinforcement.

The architecture addresses the credit-assignment problem by enabling the system to learn which actions contribute to success or failure in a complex, uncertain environment. In our study, we extend this actor-critic framework to a genomic setting by integrating established genomic metrics (e.g., the GuRu score, variant/gene risk priors, and population frequency). Instead of directly predicting variant pathogenicity, our RL method estimates the probability of observing disease-associated variants by updating weights via the temporal-difference (TD) error,

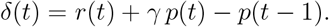

This approach quantifies the cumulative evidence that supports a variant’s clinical observability within a Bayesian context.

We systematically quantify the performance of our actor–critic RL method using simulated genomic scenarios that incorporate data imperfections such as label noise. Our goal is to identify RL methodologies and optimal parameter configurations that deliver robust, accurate predictive capabilities. These findings will form the foundation for integrating our RL framework into a broader Bayesian analytical approach for interpreting genetic evidence in clinical diagnostics.

## 2 Methods

Our investigation employed a synthetic dataset designed to mimic the characteristics of genetic variant data. The dataset comprised 2,000 variants, with 50% of the entries containing known pathogenicity labels and the remaining 50% left unlabelled for prediction purposes. Variants were generated to reflect realistic genomic scenarios by incorporating features such as the GuRu score, gene number, and population frequency. The variant distributions for each feature were designed using a mixture of priors. Variants in genes numbered 4 to 10 were assigned higher prior probabilities of pathogenicity, while variants in genes numbered 1 to 6 had lower pathogenicity priors. Genes 4–6 appeared in both categories, reflecting realistic cases where a single gene can harbour both pathogenic and benign variants depending on other genomic features. Furthermore, to simulate realistic errors and uncertainties commonly encountered in empirical genomic datasets, we intentionally introduced misannotations by randomly flipping the true pathogenicity labels for a predefined proportion (10%, 20%, and 30%) of the variants.

The GuRu score, also previously reported as the ACMGuru score, is a composite metric designed to quantify the totality of the best-known evidence regarding a genetic variant’s function, particularly its potential to be classified as pathogenic or benign. Rather than providing a direct measure of pathogenicity, the GuRu score aggregates diverse types of evidence – including clinical data, functional assays, and computational predictions – to reflect the amount of prior knowledge available about a variant. This evidence-based measure helps to objectively gauge the consensus on a variant’s classification and can be substituted by other similar metrics that integrate multiple lines of evidence, thereby offering a flexible tool for variant interpretation in genomic studies.

The reinforcement learning framework was implemented via an actor-critic algorithm. In our formulation, the state space was constructed by discretising the GuRu score into four bins, the population frequency into three bins, and the gene risk into a binary indicator; the product of these discretisations yielded 24 unique states. The action space was binary, corresponding to the two possible classifications: benign (0) or pathogenic (1). At each iteration, the RL agent observed a state corresponding to a variant and selected an action using a probabilistic policy derived from a sigmoid function applied to a vector of actor weights. A reward of +1 was granted if the predicted label matched the true pathogenicity, otherwise a penalty of -1 was imposed. The temporal-difference (TD) error, defined as the difference between the received reward and the critic’s estimate of the state value, was used to update both the actor and critic weights using learning rates *α* and *β*, respectively.

In order to assess the robustness of our model, we varied three critical parameters: the noise level in the training labels (*η*), the actor learning rate (*α*), and the critic learning rate (*β*). For each combination of these parameters, the model was trained for 20 epochs, and a variety of performance metrics were recorded.

The evaluation comprised epoch-level measures of average TD error and average reward, as well as more detailed assessments including ROC curves with associated AUC values, precision, recall, F1 scores, cumulative learning curves, and calibration plots. All these metrics were aggregated and visualised for comparison of the effects of different parameter settings.

To improve computational efficiency and allow extensive hyperparameter evaluation, we employed parallel computing through the doParallel and foreach packages in R. We used multiple cores to simultaneously evaluate different parameter combinations, reducing the computational time required for extensive simulations. Worker-specific logging, identified by process IDs, ensured clear tracking and reproducibility of parallel computations.

Synthetic data were generated according to the specified stratification and noise levels, and the RL model was trained on a randomly partitioned training set for each combination of parameters. Performance on a held-out test set was evaluated and visualised. Additionally, we generated visualisations of feature distributions, correlation matrices, and covariance matrices across noise levels, providing comprehensive diagnostic insights into the synthetic dataset’s structure and variability.

In our actor-critic RL system, the key variables are defined as follows. The actor’s weight for the *i*th connection at time *t* is denoted by *w*_*i*_(*t*) and is updated according to

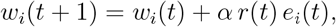

where *α* is the actor’s learning rate, *r*(*t*) is the reinforcement (or reward) signal provided by the environment at time *t*, and *e*_*i*_(*t*) is the eligibility trace for that connection. The eligibility trace *e*_*i*_(*t*) captures the recent history of activity on the *i*th connection and decays over time following

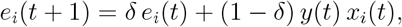

with *δ* representing the decay factor, *y*(*t*) the output of the system at time *t*, and *x*_*i*_(*t*) the *i*th component of the input vector. Meanwhile, the critic’s weight for the *i*th input is denoted by *v*_*i*_(*t*) and is updated by

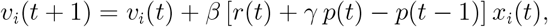

where *β* is the critic’s learning rate, *γ* is the discount factor that determines the influence of future rewards, and *p*(*t*) is the critic’s prediction of the cumulative future reward at time *t*. This architecture, by employing these update rules, addresses the credit-assignment problem by learning which actions under uncertain, delayed feedback contribute to success or failure.

## 3 Results

### 3.1 Data representation

We generated a synthetic dataset that simulates a minimally simplistic genomic variant data with both known and unknown pathogenicity. A necessary follow-up remains for the generation of highly accurate data representation and validation with real-world data. The dataset comprises key variables: GuRu Score, Population Frequency (*Pop Freq*), Gene Number (*Gene*), and ClinVar Pathogenicity (*Pathogenicity*). Known variants were produced from four distinct groups, reflecting varying profiles of these variables, while 50% of the dataset consists of variants with unknown pathogenicity, reserved for predictive modelling using reinforcement learning.

Figure 1 shows the overlaid distributions of these variables across three noise levels (0.1, 0.2, and 0.3). The figure illustrates how the distributions of *Guru Score, Pop Freq, Gene*, and *Pathogenicity* vary as the noise level increases, providing insight into the robustness of the generated data.

**Figure 1.**
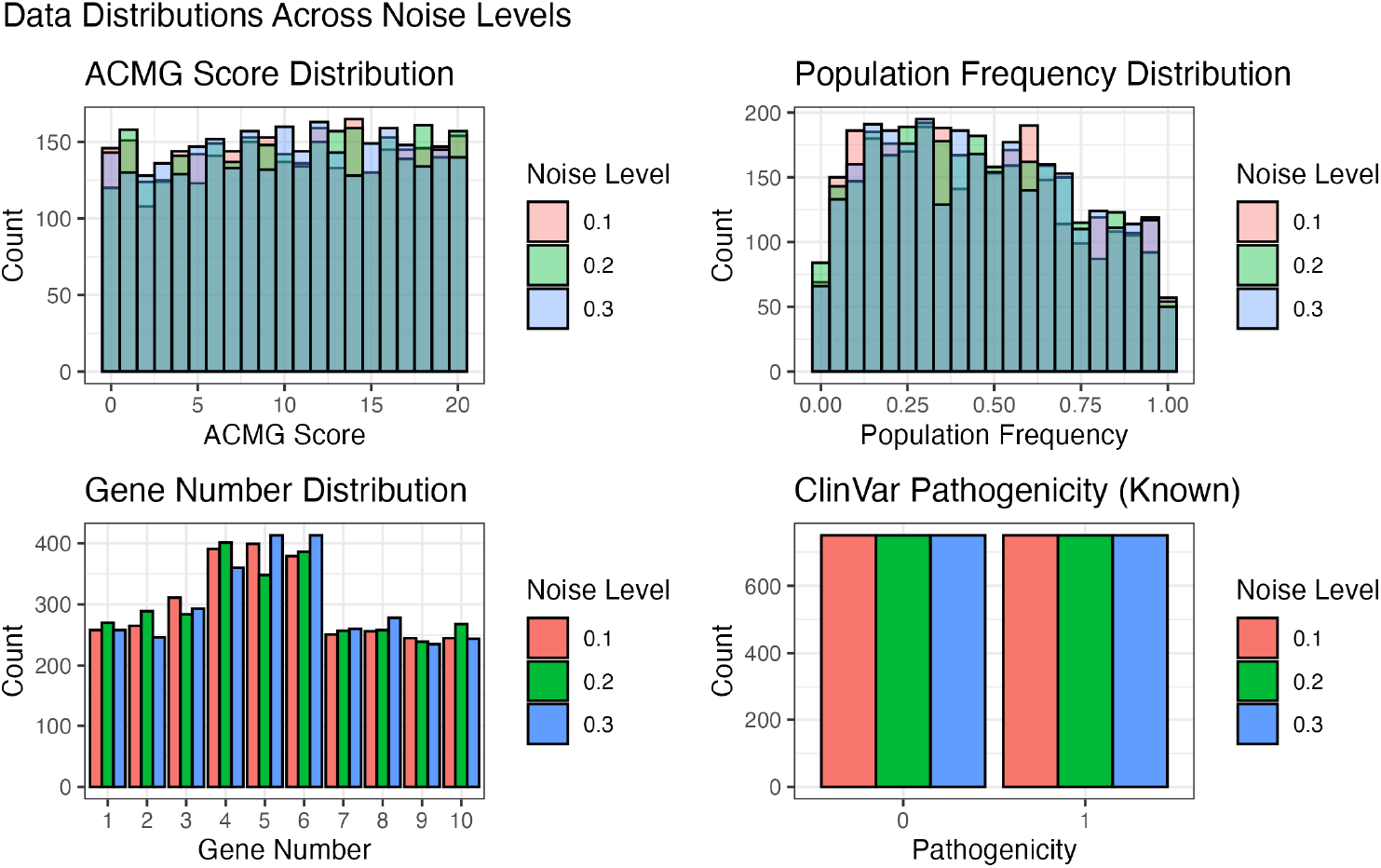
Data Distributions Across Noise Levels. This figure presents the overlaid distributions of GuRu Score, Population Frequency, Gene Number, and ClinVar Pathogenicity for noise levels 0.1, 0.2, and 0.3.

Figure 2 displays the correlation and covariance matrices for the known variants at each noise level. These matrices reveal notable relationships among the variables, such as a strong inverse correlation between *Guru Score* and *Pop Freq*, as well as a positive correlation between *Guru Score* and *Pathogenicity*. The covariance matrices further quantify the variability inherent in these relationships.

**Figure 2.**
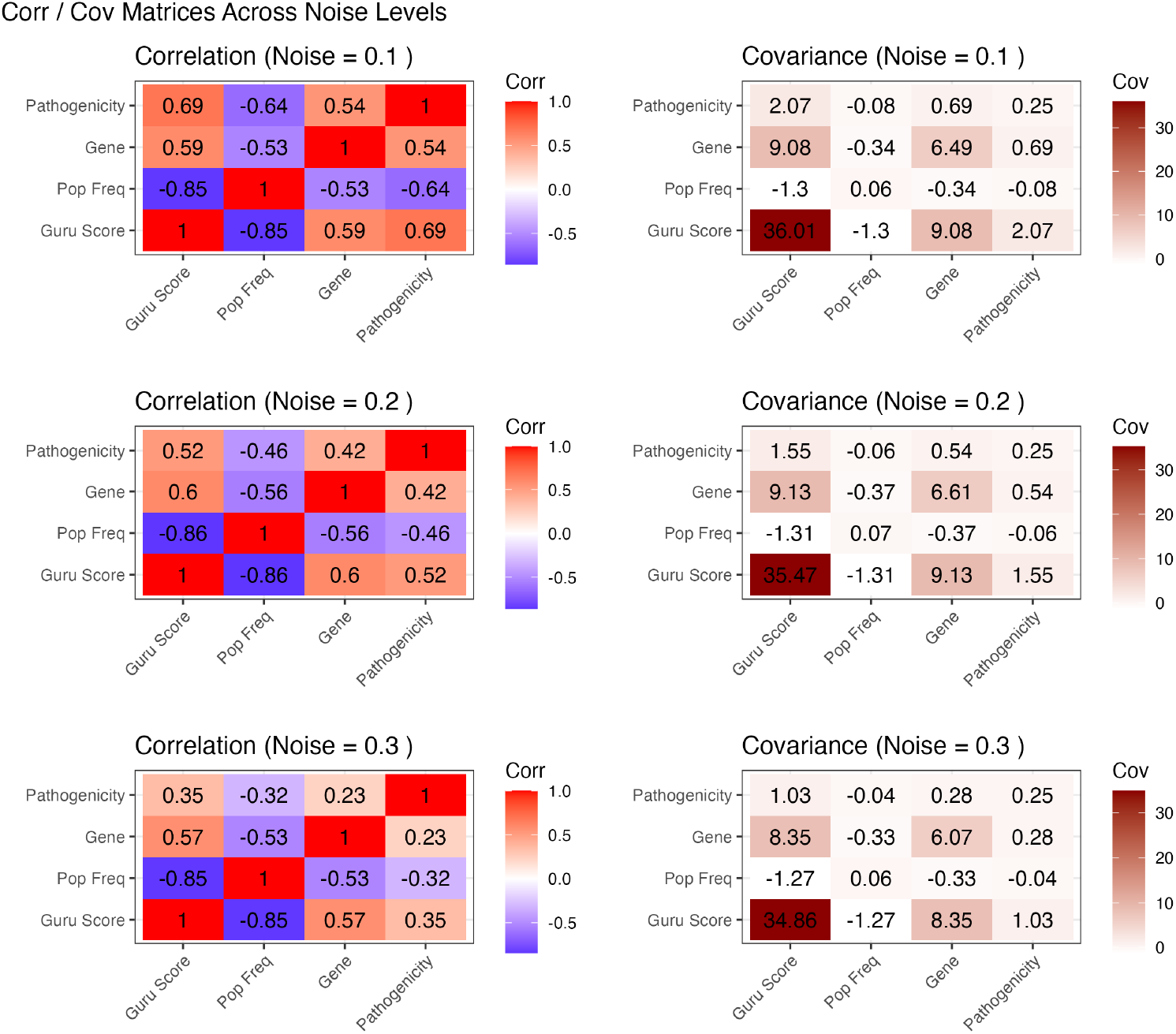
Correlation and Covariance Matrices Across Noise Levels. The figure displays the correlation (left) and covariance (right) matrices for the known variants, illustrating the relationships between *Guru Score, Pop Freq, Gene*, and *Pathogenicity* at various noise levels.

### 3.2 Performance Evaluation

In this study, we evaluated the performance of a RL model that uses GuRu scores and additional variant features to predict evidence supporting the assignment of pathogenicity in genetic data. The model was trained on known disease-causing variants and subsequently applied to predict the status of unknown variants. The experiments varied the noise level in the training labels, as well as the actor (*α*) and critic (*β*) learning rates. Performance was assessed using several metrics: average reward and TD error during training, cumulative learning curves, ROC curves with the corresponding AUC, and calibration of predicted probabilities.

Figure 3 shows the evolution of the average reward per epoch during training. This plot demonstrates that, under different parameter settings and noise levels, the model progressively improves its reward performance over successive epochs, indicating effective learning. Complementing this, Figure 4 presents the average TD error per epoch. The decreasing TD error over time reflects the convergence of the model’s value estimates and confirms that the learning algorithm is stabilising across training epochs. Figure 5 depicts the cumulative average reward over training samples, providing a continuous view of the learning progress. The gradual increase in the cumulative reward further supports the model’s ability to optimise its decision-making process over time.

**Figure 3.**
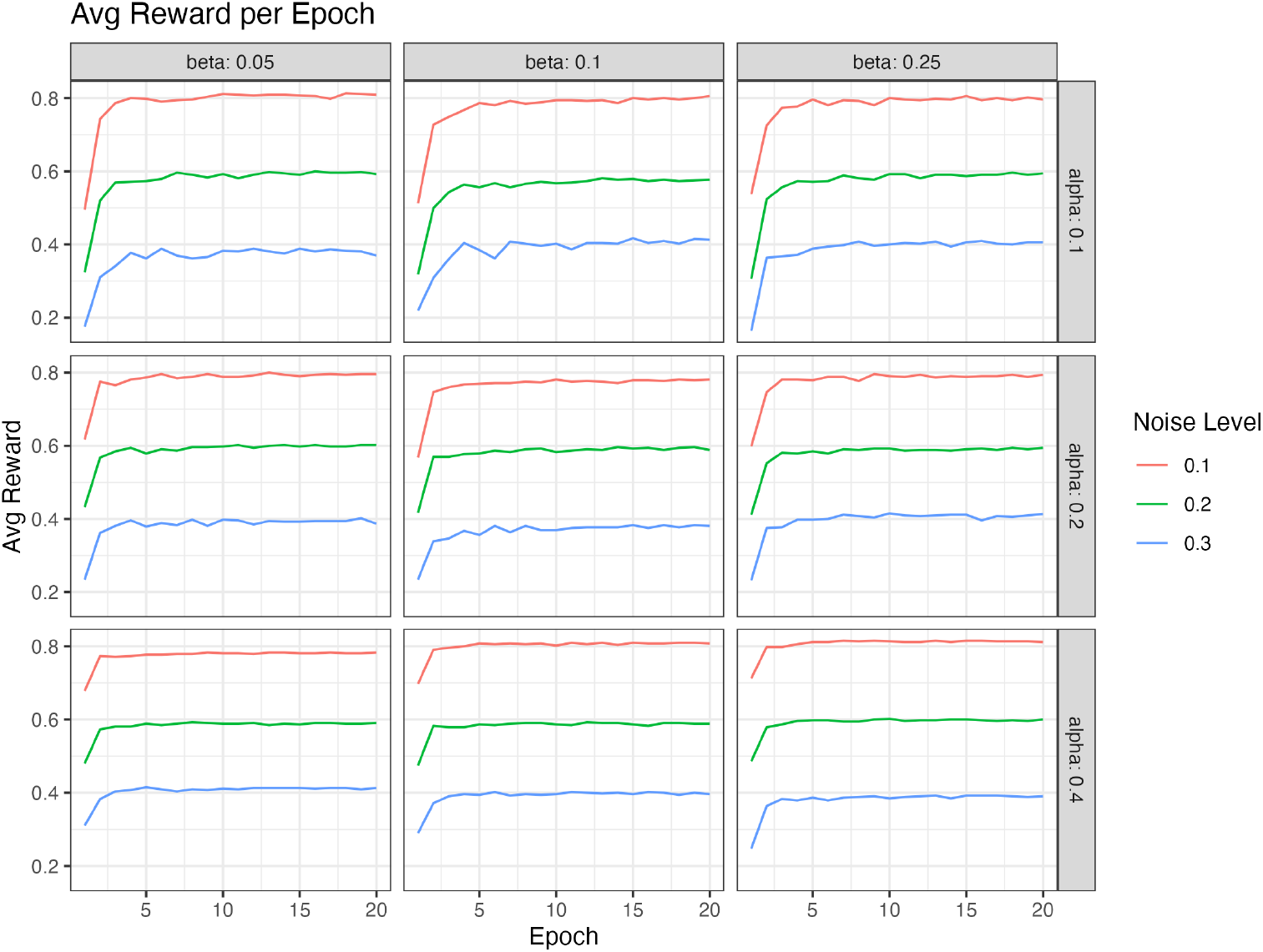
Average Reward per Epoch. This plot illustrates the evolution of the average reward during training, highlighting the model’s improving performance across different parameter settings and noise levels.

**Figure 4.**
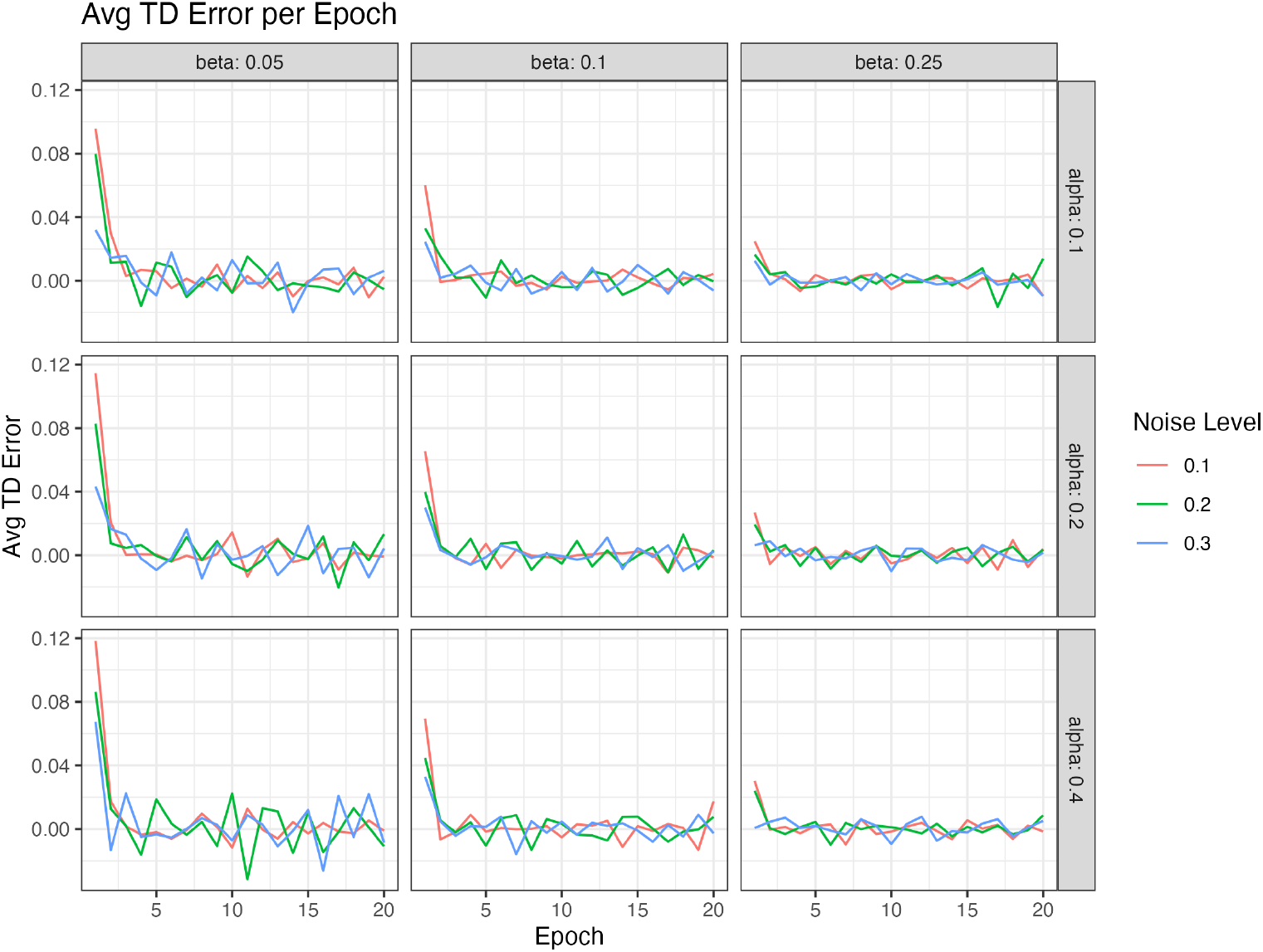
Average Temporal-Difference (TD) Error per Epoch. This figure displays the convergence behaviour of the learning algorithm, with decreasing TD error over successive epochs indicating stabilisation of the value estimates.

**Figure 5.**
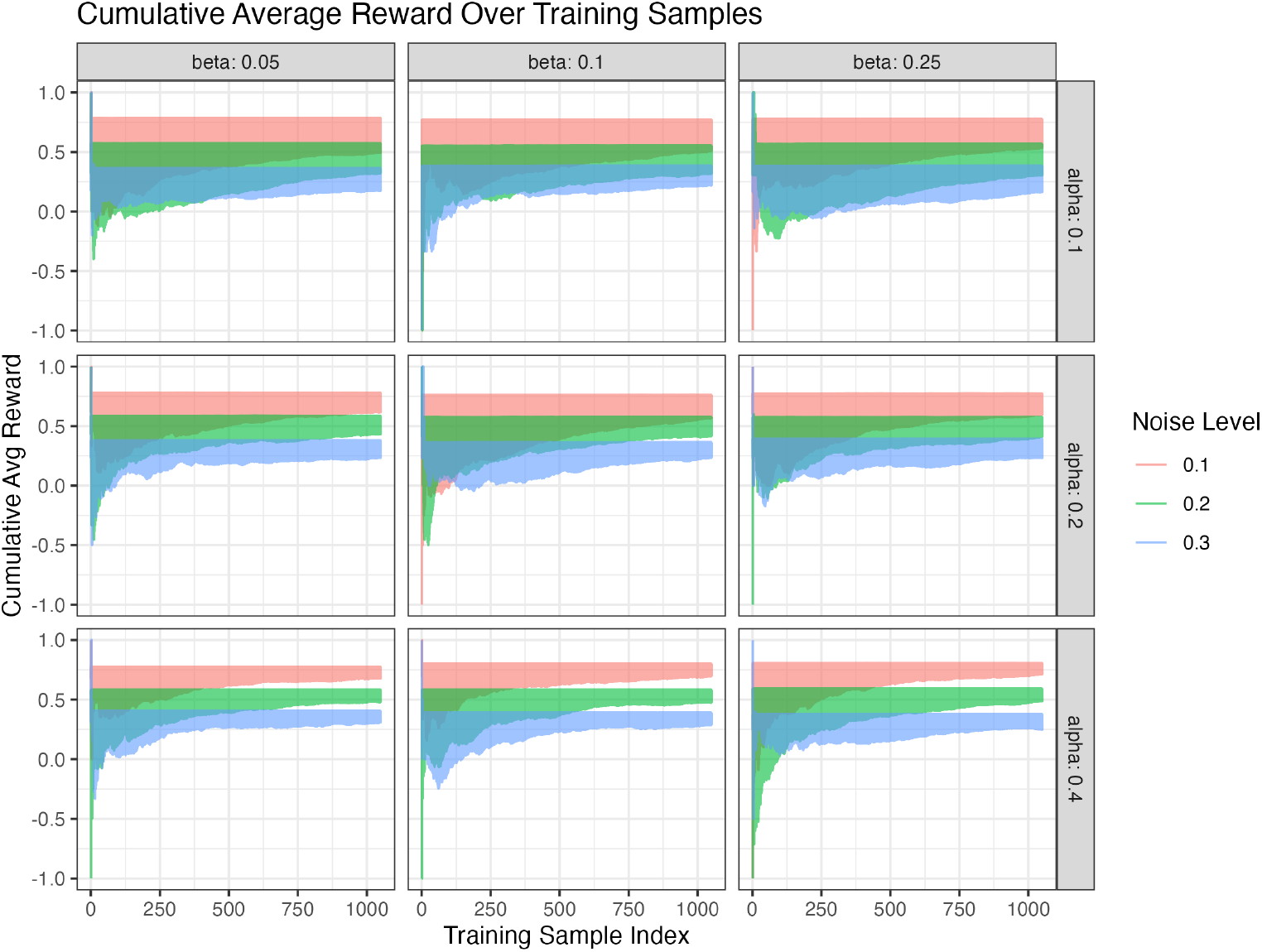
Cumulative Average Reward over Training Samples. The learning curve reflects the continuous improvement in model performance throughout training.

For evaluating the classification performance, Figure 6 displays the ROC curves for various parameter combinations. These curves illustrate the trade-off between the true positive rate and the false positive rate, with several settings yielding discrimination ability. This observation is reinforced by the AUC heatmap in Figure 7, which summarises how AUC values vary as a function of *α* and *β* for each noise level. In some cases, AUC values exceed 0.8, demonstrating adequate baseline performance.

**Figure 6.**
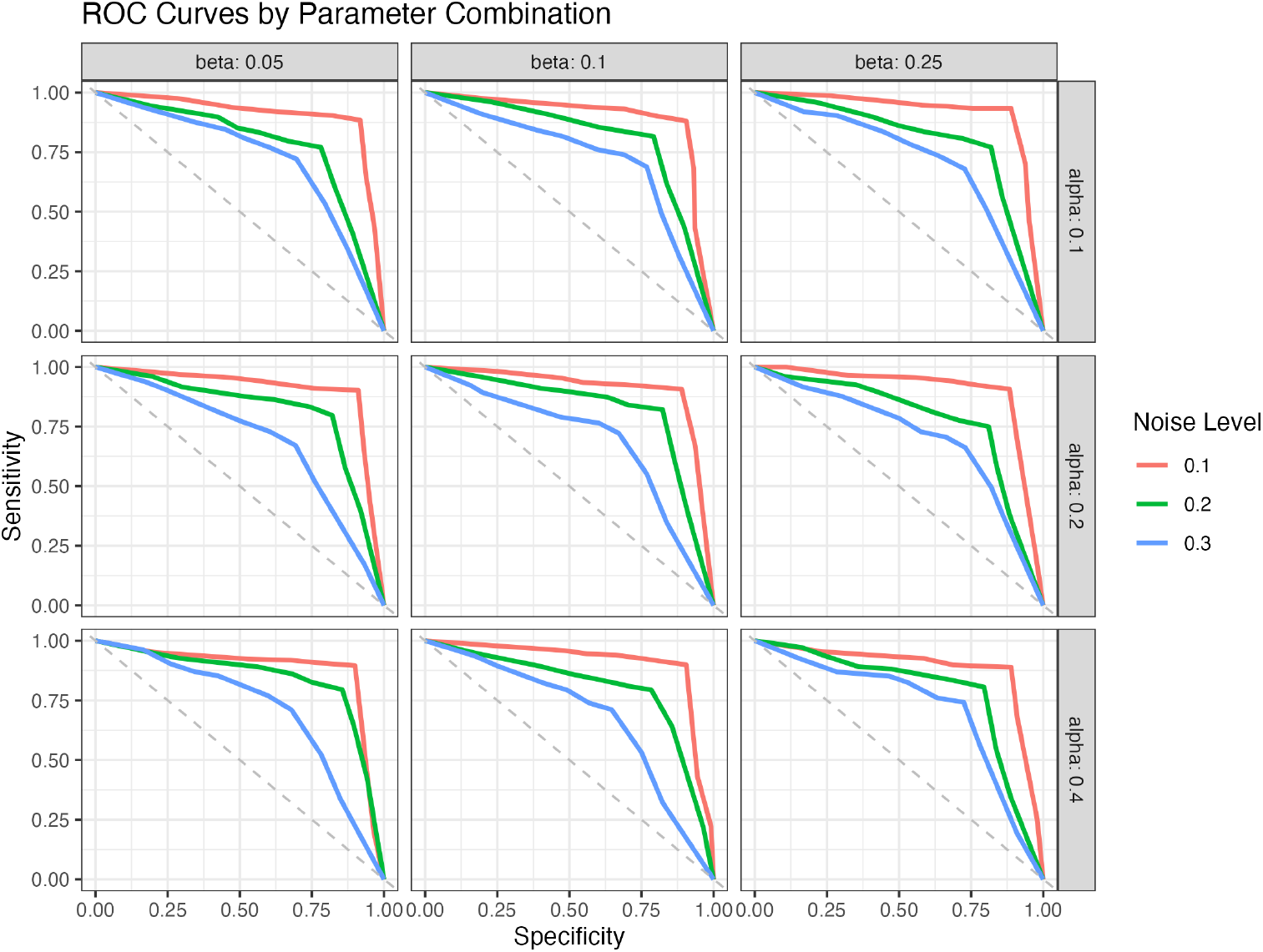
Receiver Operating Characteristic (ROC) Curves by Parameter Combination. These curves illustrate the trade-off between the true positive and false positive rates, demonstrating high discrimination ability under certain parameter settings.

**Figure 7.**
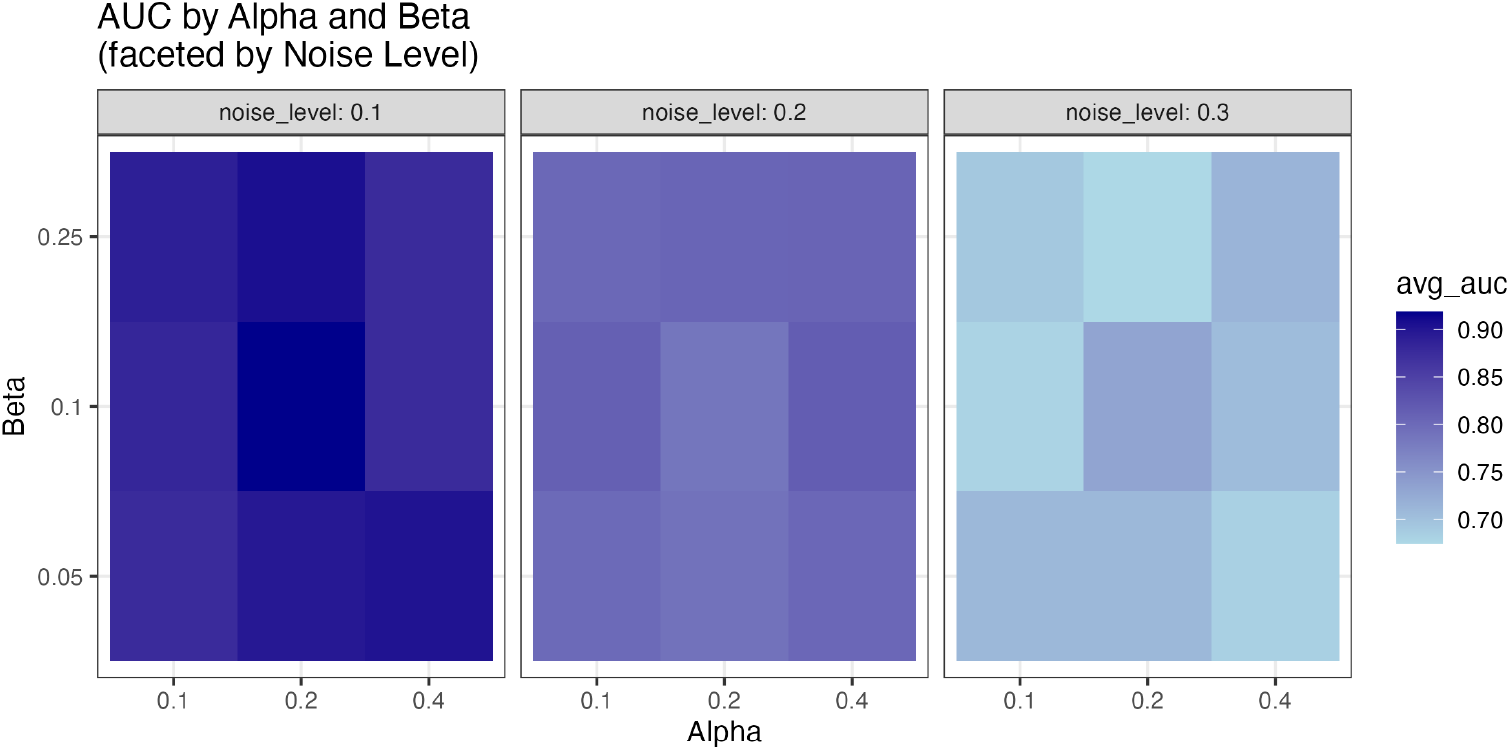
AUC Heatmap. This heatmap summarises the area under the ROC curve (AUC) as a function of the actor (*α*) and critic (*β*) learning rates, faceted by noise level. Several combinations achieve AUC values exceeding 0.8, indicating robust classification performance.

Figure 8 provides a calibration plot comparing the mean predicted probabilities to the observed proportions of pathogenic variants. Close alignment between the predicted and observed values for given parameter settings indicates that the model’s probability estimates are well-calibrated. These results demonstrate that our base-line RL framework is capable of learning to predict evidence for assigning variant pathogenicity from noisy data, with the performance being sensitive to the learning rate parameters and the level of label noise.

**Figure 8.**
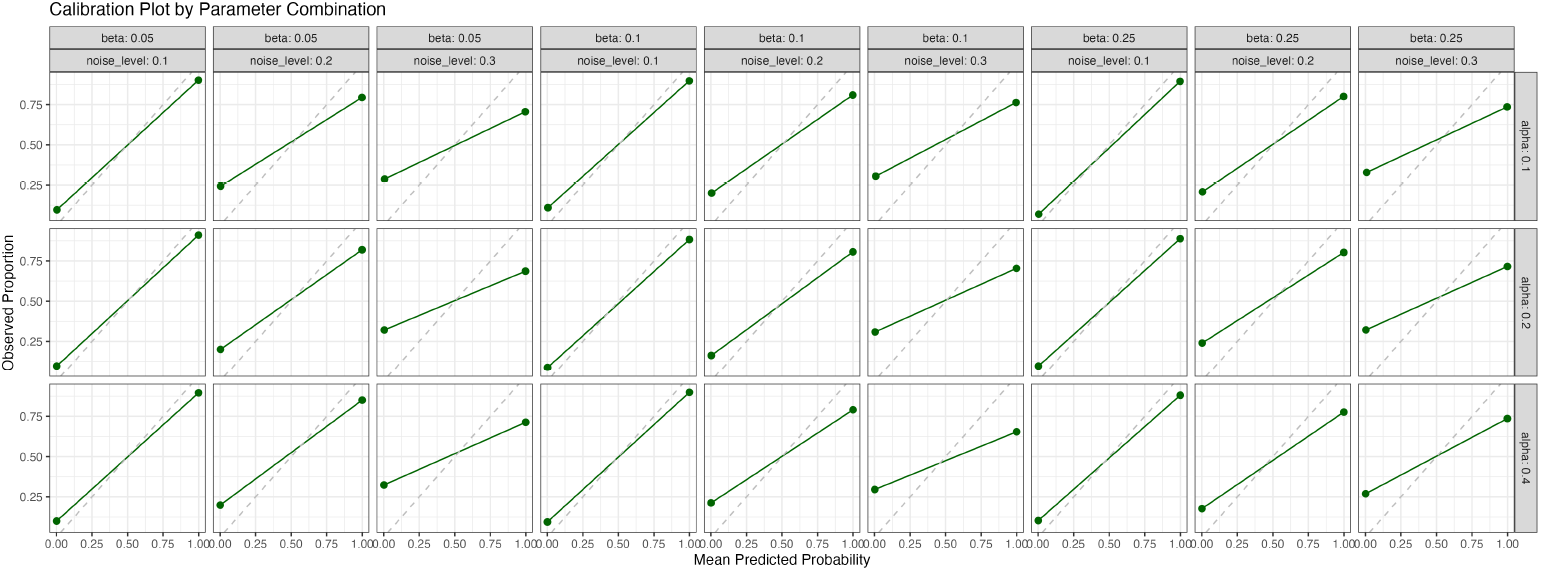
Calibration Plot. This plot compares the mean predicted probabilities to the observed proportions of pathogenic variants, showing that the predicted probabilities are well-calibrated across various parameter combinations.

## 4 Discussion

In this study, we have established an RL framework as an incremental step toward a broader Bayesian methodology aimed at classifying prior evidence about genetic variants being identified as pathogenic (or benign). Our current investigation focuses on evaluating RL methods on simulated data, explicitly quantifying multiple performance metrics relevant to the prediction of pathogenicity. By simulating scenarios reflective of real-world complexities, such as label noise and variant heterogeneity, we have demonstrated which baseline RL configurations exhibit most reliable performance.

The progressive improvement observed in average reward per epoch (Figure 3) illustrates the model’s capacity for learning despite varying levels of noise and different hyperparameter settings. Notably, the reduction in TD error across epochs (Figure 4) indicates successful convergence of our actor–critic RL algorithm. The architecture is built on two key elements: an associative search element (ASE, the actor) and an adaptive critic element (ACE, the critic) (14). The ACE, which evaluates the consequences of actions (e.g. in pole-balancing) without accessing the actor’s decisions, is adaptive-learning to predict long-term reward that, in turn, reinforces the actor’s performance. Temporal-difference learning drives this process by using successive prediction differences to reduce errors and estimate future reward in a self-supervised manner (14). Early versions estimated the gradient of the objective function stochastically, later incorporating REINFORCE to achieve stochastic gradient ascent (albeit slower than backpropagation) (15–17).

Convergence analyses of actor-critic methods remain more complex than traditional policy iteration; comprehensive results demonstrate convergence to a local maximum using a two-timescale approach (with the critic learning faster than the actor) (18).

A critical aspect of reinforcement learning is its dynamic adaptability, which we have visualised in the cumulative average reward curves (Figure 5). This upward trajectory highlights the model’s ability to progressively enhance predictive accuracy by addressing delayed reward problems-assigning appropriate credit or blame to actions long before their relevant outcomes appear. Such adaptability is imperative for practical genomic applications, where new sources of variant evidence continuously emerge, and datasets evolve.

Our analysis of ROC curves (Figure 6) and the corresponding heatmap of AUC values (Figure 7) provides a view of the predictive performance across varying parameters. Several parameter combinations produced robust discrimination performance, underscored by AUC values exceeding 0.8 even under challenging noise conditions. This finding emphasises the potential for selecting optimal RL hyperparameters tailored to specific genomic contexts and quality conditions in real datasets.

In addition to actor-critic algorithms, the 1990s witnessed the emergence of action-value methods, such as Q-learning, which map state–action pairs to expected returns and are sometimes referred to as “action-dependent adaptive critics” (19; 20). More recently, policy-gradient methods, including actor-critic algorithms, have gained traction due to their advantages in handling continuous action spaces, enabling probabilistic action selection that can converge to deterministic policies, and incorporating prior knowledge, potentially leading to faster and superior policies (21). The principles underlying these approaches have even influenced high-profile AI achievements, such as DeepMind’s Go-playing programs, which, despite their complexity, share foundational elements like dynamic environment interaction, trial-and-error search, and long-term reward prediction (14; 22–24).

Calibration analysis (Figure 8) demonstrates that the RL model can generate accurate probability estimates that closely match observed pathogenic proportions. Reliable calibration is essential for clinical interpretation, as it provides confidence in the quantitative risk assessments derived from such models. The consistency observed across a variety of parameter settings strengthens the credibility of our RL-based predictions.

This study sets the stage for subsequent application of our RL framework to real genomic data, where the complexities encountered in simulations are amplified by biological variability and data imperfections. The current simulated environment has enabled us to systematically explore and quantify algorithm performance across controlled yet realistic scenarios. Our future work will focus on applying these RL methods to empirical genomic data, further integrating Bayesian frameworks to enhance interpretability, robustness, and clinical relevance.

Ultimately, this incremental methodological development has implications for broader genomic research. By refining predictive accuracy and interpretability, RL and subsequent Bayesian methodologies hold promise for transforming evidence interpretation, enhancing clinical decision-making, and ultimately improving personalised healthcare.

## 5 Conclusion

In this study, we have developed an actor-critic RL ramework that uses the GuRu score, variant/gene risk categorisation, and population frequency to estimate the probability of observing a variant in disease, rather than directly classifying its pathogenicity. A central contribution of our work is the demonstration that RL can learn to discern which evidence supports known variant labels, thereby serving as a crucial precursor to a broader Bayesian classification framework. Our evaluations indicate that the model achieves robust predictive performance even under varying noise conditions, as evidenced by reasonable baseline AUC values, well-calibrated probability estimates, and converging learning curves. These findings pave the way for future integration of RL with Bayesian inference in clinical genomics, ultimately enhancing the accuracy and genetic variant interpretation and improving personalized healthcare outcomes.

## Data Availability

All data produced are available online at https://github.com/DylanLawless/rl2025lawless

## Acronyms

**ACE** associative search element

**ASE** adaptive critic element

**AUC** area under the curve

**ROC** receiver operating characteristic

**QV** Qualifying variant

**RL** reinforcement learning

**TD** temporal-difference

